# Performance characteristics of a high throughput automated transcription mediated amplification test for SARS-CoV-2 detection

**DOI:** 10.1101/2020.07.06.20143719

**Authors:** Jimmykim Pham, Sarah Meyer, Catherine Nguyen, Analee Williams, Melissa Hunsicker, Ian McHardy, Inessa Gendlina, D. Yitzchak Goldstein, Amy S. Fox, Angela Hudson, Paul Darby, Paul Hovey, Jose Morales, James Mitchell, Karen Harrington, Mehrdad Majlessi, Joshua Moberly, Ankur Shah, Andrew Worlock, Marion Walcher, Barbara Eaton, Damon Getman, Craig Clark

## Abstract

The COVID-19 pandemic caused by the new SARS-CoV-2 coronavirus has imposed severe challenges on laboratories in their effort to achieve sufficient diagnostic testing capability for identifying infected individuals. In this study we report the analytical and clinical performance characteristics of a new, high-throughput, fully automated nucleic acid amplification test system for the detection of SARS-CoV-2. The assay utilizes target capture, transcription mediated amplification, and acridinium ester-labeled probe chemistry on the automated Panther System to directly amplify and detect two separate target sequences in the ORF1ab region of the SARS-CoV-2 RNA genome. The probit 95% limit of detection of the assay was determined to be 0.004 TCID_50_/ml using inactivated virus, and 25 c/ml using synthetic *in vitro* transcript RNA targets. Analytical sensitivity (100% detection) was confirmed to be 83 – 194 c/ml using three commercially available SARS-CoV-2 nucleic acid controls. No cross reactivity or interference was observed with testing six related human coronaviruses, as well as 24 other viral, fungal, and bacterial pathogens, at high titer. Clinical nasopharyngeal swab specimen testing (N=140) showed 100%, 98.7%, and 99.3% positive, negative, and overall agreement, respectively, with a validated reverse transcription PCR NAAT for SARS-CoV-2 RNA. These results provide validation evidence for a sensitive and specific method for pandemic-scale automated molecular diagnostic testing for SARS-CoV-2.

## INTRODUCTION

Coronavirus disease-19 (COVID19) is a novel respiratory illness caused by severe acute respiratory syndrome coronavirus 2 (SARS-CoV-2), a novel *Sarbecovirus* that emerged from the region of Wuhan, China in late 2019 (1). People with COVID-19 experience mild to severe respiratory symptoms including fever, cough and shortness of breath or difficulty breathing (2), although some individuals experience no symptoms at all (3).

The COVID-19 pandemic has occurred across all continents, adding more than 100,000 new SARS-CoV-2 cases globally each day (4,5). As communities begin reopening and relaxing quarantine measures, there is the potential risk for an upsurge in cases and rates of viral transmission. The availability of validated high-throughput diagnostic tests is therefore essential for rapidly and efficiently informing patient management decisions, implementing hospital infection prevention practices, and for guiding public health responses to wide-scale infection control measures to reduce transmission in populations.

To meet the need for pandemic-scale diagnostic testing, we have developed and validated a high-throughput, fully automated nucleic acid amplification test (NAAT) for direct amplification and detection of SARS-CoV-2 RNA from specimens of infected individuals. The assay employs target capture, transcription mediated amplification (TMA) and acridinium ester-labeled probe chemistries to enable a sample-to-result solution for detection of two different conserved target regions within the ORF1ab region of the SARS-CoV-2 genome. Herein, we describe the analytical and clinical performance characteristics of the assay.

## MATERIALS AND METHODS

### Transcription Mediated Amplification Test for SARS-CoV-2

The Aptima® SARS-CoV-2 assay utilizes magnetic bead-based target capture, isothermal TMA of RNA, and dual kinetic acridinium ester-labeled probe hybridization for the isolation, amplification, and detection of an internal process control RNA, and two unique sequences within the ORF1ab region of the SARS-CoV-2 viral genome. The assay is performed on the automated Panther® and Panther Fusion® instruments (both from Hologic Inc, San Diego, USA), and received FDA Emergency Use Authorization (EUA) on May 14, 2020. It is intended for the qualitative detection of SARS-CoV-2 RNA isolated and purified from nasopharyngeal (NP) swab, nasal swab (NS), mid-turbinate and oropharyngeal (OP) swab, NP wash/aspirate, or nasal aspirate specimens, obtained from individuals meeting COVID-19 clinical and/or epidemiological criteria. Sample input volume is 0.5 ml, with continuous sample and reagent loading access, automated RNA extraction, amplification, detection, and results reporting. Time to first result is 3 h 30 min, with a capacity of approximately 1,025 results per 24 h per instrument system.

### Comparison Assay

The validated EUA Panther Fusion SARS-CoV-2 reverse transcription PCR (RT-PCR) assay (Hologic Inc.) was used as a comparator assay for clinical performance studies. This assay was performed as previously described (6).

### Analytical Performance

#### Limit of Detection

The analytical sensitivity of the SARS-CoV-2 TMA assay was assessed using 2 lots of reagents to test 60 replicates each of dilution panels containing cultured SARS-CoV-2 virus strain USA-WA1/2020 (BEI Resources, Manassas, VA) and diluted in Aptima specimen transport medium (STM) matrix to a range of 0.03 to 0.0003 tissue culture infectious dose 50 per ml (TCID_50_/ml). Also tested were replicates (n = 43 to 60) of panels consisting of two *in vitro* transcribed (IVT) RNA targets, corresponding to two unique target sequences within the ORF1ab region of the SARS-CoV-2 RNA genome, diluted in STM. Assay positivity for both studies was determined using a pre-defined cutoff value of 560 kilo relative light units (kRLU). Results were analyzed by probit analysis (normal model) to determine the 95% limit of detection (LOD). Analytical sensitivity was confirmed by testing 20 replicates each of SARS-CoV-2 virus diluted in four specimen matrices (pooled NP swab, STM, saline and Liquid Amies transport medium (Copan, Murrieta, CA)) at 0.003 TCID_50_/ml for NP swab, STM and saline, and 0.003 and 0.01 TCID_50_/ml for Liquid Amies.

#### Analytical Specificity/Interference

Analytical specificity of the SARS-CoV-2 TMA assay was determined by evaluating assay cross-reactivity and interference using 30 non-target microorganisms (17 viral species, including 6 non - SARS-CoV-2 coronaviruses, 11 bacterial species, and 2 fungal species; N=3 replicates each) at the highest titer achievable. Thirty NP swab specimens obtained from consented asymptomatic donors were also tested to represent diverse microbial flora in the human respiratory tract. Interference by high-titer non-target organisms was assessed by testing of 3 replicates of each organism in the presence of low titer (0.03 TCID_50_/ml) SARS-CoV-2 virus. Cross reactivity by high-titer non-target organisms was assessed in the absence of SARS-CoV-2 virus.

#### SARS-CoV-2 Commercial Control Panel Testing

Stock SARS-CoV-2 control panel materials from three commercial suppliers (Exact Diagnostics (Fort Worth, TX) SARS-CoV-2 Standard, cat no. COV019; SeraCare (Milford, MA) AccuPlex SARS-CoV-2 Verification Panel, cat no. 0505-0126; and ZeptoMetrix (Buffalo, NY) SARS-CoV-2 External Run Control cat no. NATSARS(COV2)-ERC0831042) were diluted in STM to 6 concentrations ranging from 833 to 8 copies per ml (c/ml) and multiple replicates (n = 20 to 40) were tested with the SARS-CoV-2 TMA assay. The control panels were also tested with the SARS-CoV-2 RT-PCR assay on the automated Panther Fusion platform.

### Clinical Performance

#### Specimen Collection

Residual de-identified NP swab samples were collected by standard methods from 140 symptomatic patients at two different US clinical sites (San Diego, CA and The Bronx, NY). Specimens were transported to the laboratory and tested with the SARS-CoV-2 TMA assay and the Fusion SARS CoV-2 RT-PCR assay. An additional clinical sample set consisting of paired NP swab, OP swab and NS specimens were collected from 38 patients; complete sets (containing all three specimen types) were obtained from 35 patients. NS samples were collected first by inserting the swab into the subject’s nostril past the inferior turbinate, approximately 3 cm, twisting the swab in mid-turbinate area for 3 to 5 seconds and placing the swab into a tube of STM. OP swab samples were collected immediately following NS samples by swabbing the posterior pharynx for 3-5 seconds and placing the swab into a specimen tube. Samples were frozen and shipped to Hologic (San Diego, CA) for testing.

## RESULTS

Data for the analytical sensitivity determination of the SARS-CoV-2 TMA assay are shown in **Figure 1**. Using a pre-determined cutoff value of 560 kRLU, the assay yielded 100% positivity at a concentration of 0.01 TCID_50_/ml of SARS-CoV-2 virus and at 100 c/ml of SARS-CoV-2 IVT RNA targets. Using probit analysis, the 95% limit of detection was determined to be 0.004 TCID_50_/ml (95% CI: 0.003 – 0.007) for SARS-CoV-2 virus, and 25.4 c/ml (95% CI: 16.9 − 50.5) for ORF1ab IVT RNA targets. Analytical sensitivity for the assay was confirmed by testing SARS-CoV-2 virus in 4 specimen matrices (pooled NP swab specimens, STM, saline, and Liquid Amies transport medium) at 0.003 TCID_50_/ml. All specimen matrices yielded 95% positivity (19/20 replicates detected) at this concentration, except Liquid Amies transport medium, which was 85% (17/20) positive at 0.003 TCID_50_/ml and 100% (20/20) positive at 0.01 TCID_50_/ml (**Table 1**).

**Figure 1.**
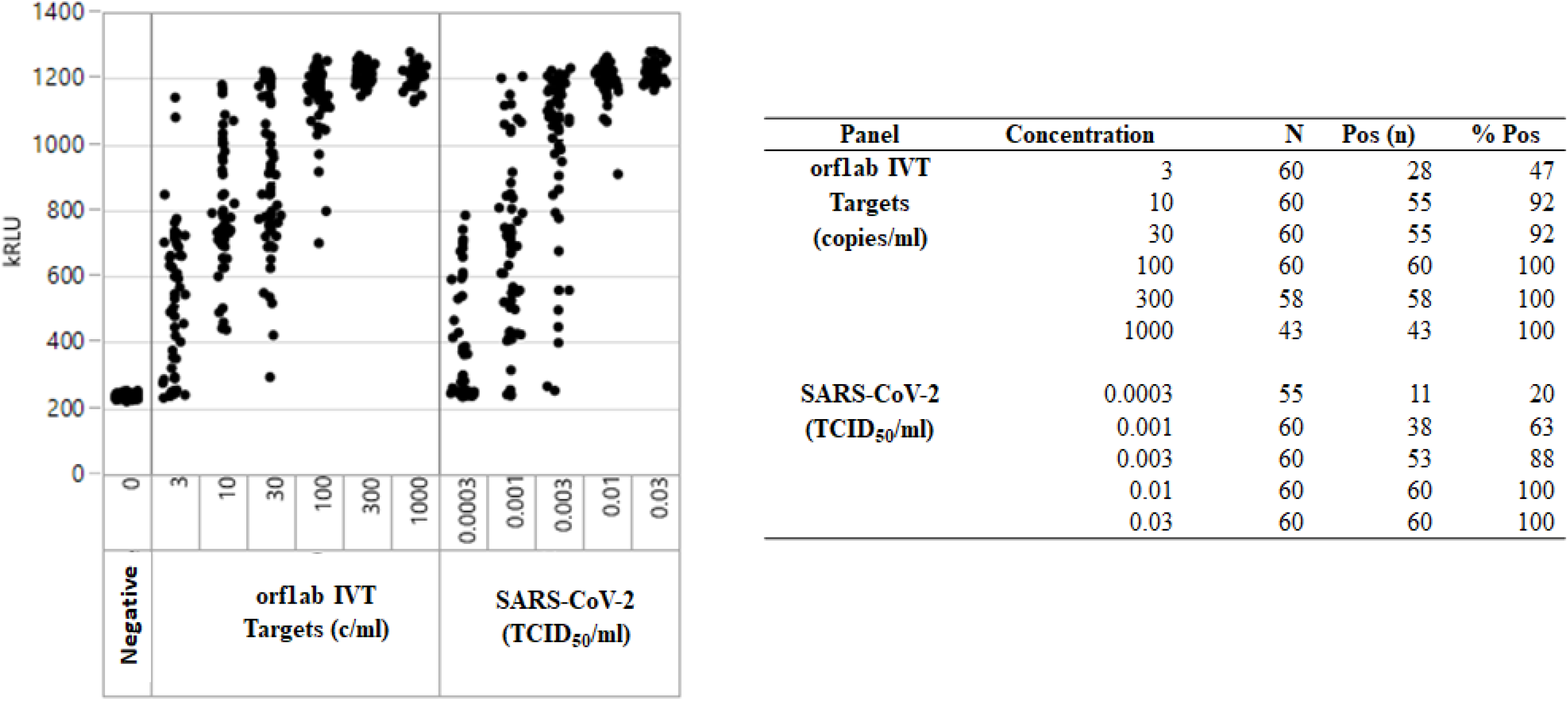
Analytical sensitivity of automated Aptima SARS-CoV-2 TMA assay for detection of SARS-CoV-2 Open Reading Frame 1ab (orf1ab) RNA. IVT, *in vitro* RNA transcript; TCID_50_, Tissue Culture Infectious Dose 50%.

**Table 1.** Confirmation of Aptima SARS-CoV-2 TMA assay limit of detection in different specimen matrices.

| Target | Matrix | TCID<sub>50</sub>/ml | n/N (%) | Average kRLU (%CV) |
| --- | --- | --- | --- | --- |
| SARS-CoV-2 virus | NP | 0 | 0/0 (0) | 278 (2.9) |
|  |  | 0.003 | 19/20 (95) | 908 (17.2) |
|  | STM | 0 | 0/0 (0) | 289 (2.2) |
|  |  | 0.003 | 19/20 (95) | 835 (24.4) |
|  | Saline | 0 | 0/0 (0) | 288 (2.4) |
|  |  | 0.003 | 19/20 (95) | 876 (24.8) |
|  | Liquid Amies | 0 | 0/0 (0) | 286 (1.8) |
|  |  | 0.003 | 17/20 (85) | 877 (24.7) |
|  |  | 0.01 | 20/20 (100) | 1100 (3.9) |

STM, Aptima specimen transport medium. kRLU, kilo relative light unit.

Analytical specificity of the SARS-CoV-2 TMA assay was determined by evaluating assay cross-reactivity and interference using 30 non-target viral, bacterial and fungal microorganisms at the highest titer achievable, as well as in 30 NP swab specimens obtained from consented asymptomatic donors at low risk of SARS-CoV-2 infection. As shown in the **Supplemental Table**, none of the microorganisms or NP swab specimens tested caused cross-reactivity in the absence of SARS-CoV-2 target or interfered with TMA detection in the presence of SARS-CoV-2 spiked at 0.03 TCID_50_/ml.

The clinical accuracy of the SARS-CoV-2 TMA assay was compared to the SARS-CoV-2 RT-PCR assay using 140 patient NP swab specimens (**Table 2**). This analysis resulted in positive, negative, and overall agreements of 100% (95%CI: 94.3% - 100%), 98.7% (95%CI: 92.9% - 99.8%), and 99.3% (95%CI: 96.1% - 99.3%), respectively. Clinical performance of the SARS-CoV-2 TMA assay was also assessed by testing sets of NP swabs, OP swabs, and nasal swabs co-collected from 35 symptomatic patients suspected of being infected with SARS-CoV-2. **Figure 2** shows the SARS-CoV-2 TMA assay had 100% positive and negative agreements of the NP swab specimens with the co-collected OP swab and nasal swab specimens. Similar results were obtained for the paired specimen sets using the SARS-CoV-2 RT-PCR assay (**Supplemental Figure**).

**Table 2.**
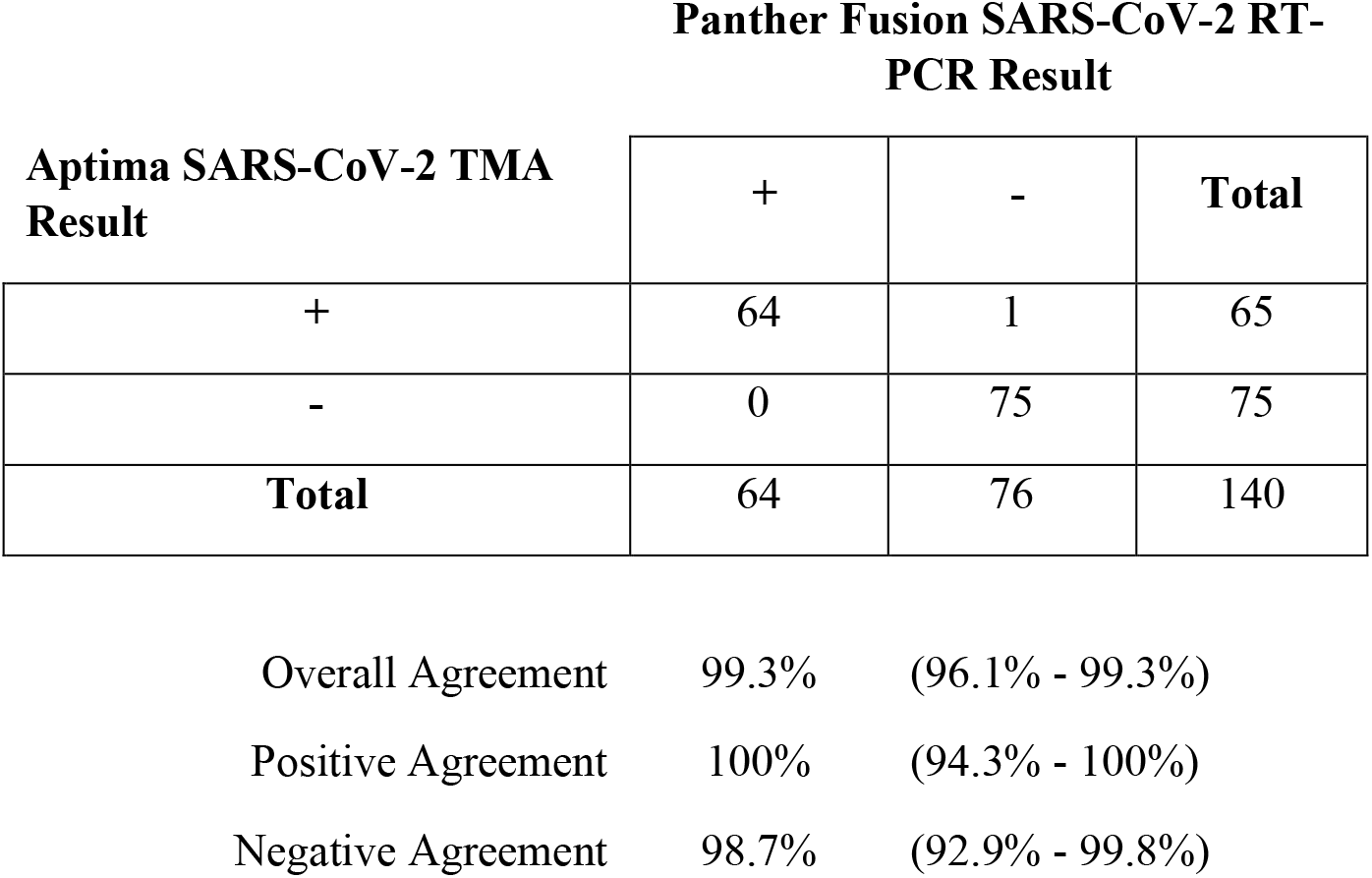
Agreement analysis (%, (95% confidence interval) between the Aptima SARS-CoV-2 TMA assay and the Panther Fusion SARS-CoV-2 RT-PCR assay for nasopharyngeal swab specimens.

| **Aptima SARS-CoV-2 TMA**<br>**Result** | **Panther Fusion SARS-CoV-2 RT-**<br>**PCR Result** |  |  |
| --- | --- | --- | --- |
|  | **+** | **-** | **Total** |
| **+** | 64 | 1 | 65 |
| **-** | 0 | 75 | 75 |
| **Total** | 64 | 76 | 140 |

|  |  |  |
| --- | --- | --- |
| Overall Agreement | 99.3% | (96.1% - 99.3%) |
| Positive Agreement | 100% | (94.3% - 100%) |
| Negative Agreement | 98.7% | (92.9% - 99.8%) |

**Figure 2.**
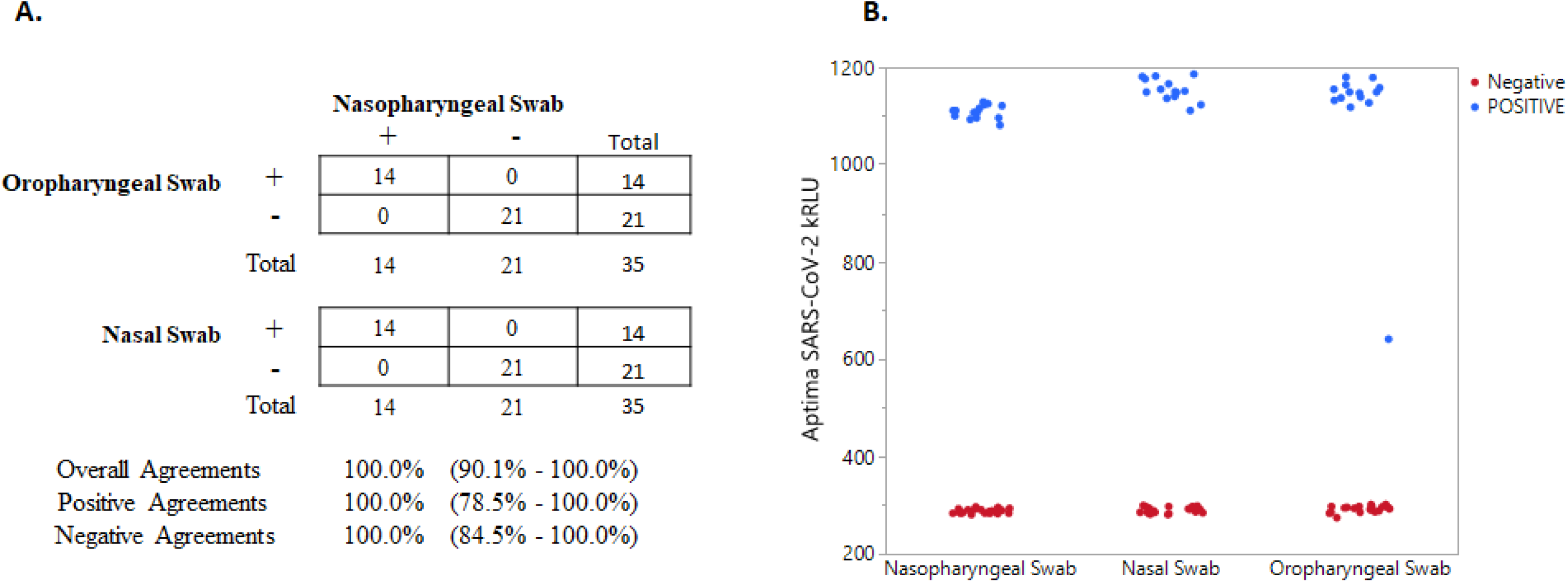
Agreement (**A**) between 35 sets of co-collected nasopharyngeal swab, oropharyngeal swab, and nasal swab clinical specimens with positive (+) and negative (-) Aptima SARS-CoV-2 TMA assay results. Scatter plot (**B**) of corresponding Aptima SARS-CoV-2 TMA assay positive and negative kRLU signal for each sample type.

SARS-CoV-2 control panel materials from three commercial suppliers (Exact Diagnostics, SeraCare, ZeptoMetrix) were evaluated by building dilution panels of each control material and testing multiple replicates with both the SARS-CoV-2 TMA assay and the SARS-CoV-2 RT-PCR assay (**Table 3**). Both assays yielded similar results. For the Exact Diagnostics SARS-CoV-2 control, the TMA and RT-PCR assays each had 100% detection down to 83 c/ml (N=40 each). For the SeraCare SARS-CoV-2 control, the TMA assay was 100% at 83 c/ml (N=20) while the RT-PCR assay was 90% at 83 c/ml and 100% at 194 c/ml (N=20 for both). The SARS-CoV-2 TMA assay (N=37) and the SARS-CoV-2 RT-PCR assay (N=40) were both 100% reactive at 194 c/ml using the ZeptoMetrix control material.

**Table 3.** Performance of Aptima SARS-CoV-2 TMA and Panther Fusion SARS-CoV-2 RT-PCR assays for detection of commercially available SARS-CoV-2 controls.

| **SARS-CoV-2 Control Vendor** | **Concentration (c/ml)** | **Aptima SARS-CoV-2 TMA n/N (%)** | **Panther Fusion SARS-CoV-2 RT-PCR n/N (%)** |
| --- | --- | --- | --- |
| Exact Diagnostics SARS-CoV-2 Standard | 833 | 40/40 (100) | 38/38 (100) |
|  | 417 | 40/40 (100) | 39/39 (100) |
|  | 194 | 40/40 (100) | 40/40 (100) |
|  | 83 | **40/40 (100)** | **40/40 (100)** |
|  | 19 | 35/39 (90) | 30/40 (75) |
|  | 8 | 29/39 (74) | 21/40 (53) |
| SeraCare Accuplex™ SARS-CoV-2 Verification Panel | 833 | 20/20 (100) | 20/20 (100) |
|  | 417 | 19/19 (100) | 20/20 (100) |
|  | 194 | 19/19 (100) | **20/20 (100)** |
|  | 83 | **20/20 (100)** | 18/20 (90) |
|  | 19 | 16/40 (40) | 11/40 (28) |
|  | 8 | 7/40 (18) | 4/40 (10) |
| ZeptoMetrics SARS-CoV-2 (recombinant) | 833 | 39/39 (100) | 40/40 (100) |
|  | 417 | 40/40 (100) | 40/40 (100) |
|  | 194 | **37/37 (100)** | **40/40 (100)** |
|  | 83 | 39/40 (97.5) | 32/40 (80) |
|  | 19 | 12/40 (30) | 14/40 (35) |
|  | 8 | 12/40 (30) | 8/40 (20) |

## DISCUSSION

The availability of large-scale diagnostic testing for SARS-CoV-2 addresses a current critical need for identifying the prevalence and spread of the virus in populations and for guiding public health policies and interventions to minimize incident infections. This study describes the performance characteristics of a new high-throughput isothermal TMA NAAT that employs a complete sample-to-result automation system to maximize sample testing throughput in clinical laboratories. The analytical performance data demonstrates the assay is highly sensitive and specific for detection of viral SARS-CoV-2 RNA and has high clinical agreement (100% positive agreement, 98.7% negative agreement) with a EUA validated RT-PCR assay for SARS-CoV-2 RNA.

The unprecedented need for SARS-CoV-2 testing has resulted in national shortages in sample collection materials including VTM, prompting the CDC and FDA to recommend optional specimen collection media, such as saline and Liquid Amies (7, 8). To ensure comparable performance across various collection media, we evaluated analytical sensitivity of the SARS-CoV-2 TMA assay in NP swab matrix (NP swabs collected in VTM), STM, saline, and Liquid Amies transport medium using inactivated SARS-CoV-2 virus. NP swab matrix, Aptima STM, and saline demonstrated 95% detection at 0.003 TCID_50_/ml, whereas Liquid Amies showed slightly lower sensitivity at 0.01 TCID_50_/ml. Despite this small difference in analytical sensitivity, these results suggest that all media are acceptable for use for clinical sample collection.

NP swab specimens have been considered the gold standard sampling method for respiratory virus infection (9) as it has been reported that nasal swab or OP swab samples may have a slightly lower sensitivity compared to NP swabs (10, 11). However, given the challenges of collection device shortages for SARS-CoV-2 diagnosis, particularly NP swabs and VTM, the use of nasal swab and OP swab as alternate samples for diagnosis of SARS-CoV-2 has been evaluated. The CDC recently removed NP swab as the “preferred” sample type from their sample collection guidelines (7) and others have reported comparable performance of NS and OP swabs for the diagnosis of SARS-CoV-2 (12-15). Our data shows strong agreement between SARS-CoV-2 detection from NS and OP swabs compared to paired NP swab samples for the SARS-CoV-2 TMA and SARS-CoV-2 RT-PCR assays. We did observe for one patient positive NP and nasal swab results, but negative results in the paired OP swab, however overall the data indicate that both NS and OP swab specimens are adequate sample types for diagnosis of SARS-CoV-2.

The performance of other SARS-CoV-2 NAATs that have received Emergency Use Authorization has been characterized using commercially available inactivated virus preparations (e.g., BEI Resources, Manassas, VA) or synthetic RNA quality control materials. Our evaluation of three different external RNA control materials demonstrated comparable analytical sensitivity with both the SARS-CoV-2 TMA assay and the SARS-CoV-2 RT-PCR assay, with LOD values between 83 c/ml – 194 c/ml. These analytical sensitivity values correlate with previously reported LOD values by Zhen *et al* (16). However, using LOD values as determined by external control material to assess or compare assay performance warrants some caution as different control materials may give considerably different results for the absolute LOD value (16) and the reported RNA or DNA stock concentrations of these materials may differ from true concentrations (17).

Limitations of this study include the small number of clinical specimens available for testing, and the absence of discordant result resolution by testing with a third assay. The paired specimen testing (NS and OP) included only 35 patients (14 of which were positive). The specimens that were included did span a typical clinical titer range for the virus (SARS-CoV-2 RT-PCR Ct values ranged from 14.5 to 37.1). Additional clinical data should be collected to more fully assess and compare performance between these sample types.

In summary, the SARS-CoV-2 TMA assay is highly sensitive and specific and provides an automated high-throughput testing solution for large-scale diagnostic testing for the virus. The assay system is able to process and generate results for > 1,000 samples per day enabling medical centers, reference laboratories and public health laboratories to efficiently process and analyze very high volumes of specimens for the detection of SARS-CoV-2 RNA (18).

## Data Availability

All data are available for review upon request

---

**Supplemental Figure.**
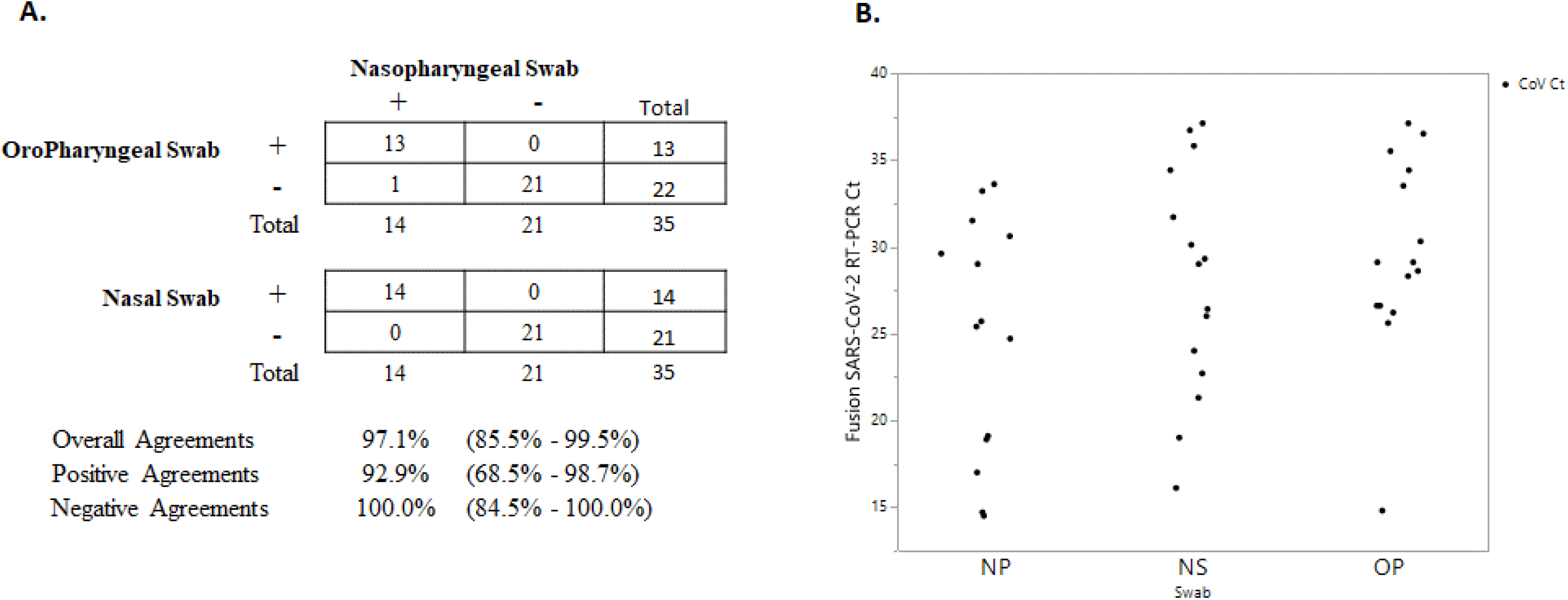
Agreement (**A**) between 35 sets of co-collected nasopharyngeal swab, oropharyngeal swab, and nasal swab clinical specimens with positive (+) and negative (-) Panther Fusion SARS-CoV-2 RT-PCR assay results. Scatter plot (**B**) of RT-PCR Ct values corresponding to Panther Fusion SARS-CoV-2 RT-PCR assay positive samples for each swab type.

**Supplemental Table.** Microorganism cross reactivity and interference of the Aptima SARS-CoV-2 TMA assay.

| Microorganism | Concentration | SARS-CoV-2<br>Unspiked |  |  | SARS-CoV-2<br>Spiked <sup>a</sup> |  |  |
| --- | --- | --- | --- | --- | --- | --- | --- |
|  |  | N Tested | N Detected | % Detected | N Tested | N Detected | % Detected |
| No organism Control | N/A | 3 | 0 | 0 | 3 | 3 | 100 |
| Human coronavirus 229E | 1.00E+05 TCID50/ml | 3 | 0 | 0 | 3 | 3 | 100 |
| Human coronavirus OC43 | 1.00E+05 TCID50/ml | 3 | 0 | 0 | 3 | 3 | 100 |
| Human coronavirus HKU1 | 1.00E+06 c/ml | 3 | 0 | 0 | 3 | 3 | 100 |
| Human coronavirus NL63 | 1.00E+04 TCID50/ml | 3 | 0 | 0 | 3 | 3 | 100 |
| SARS-coronavirus | 1.00E+06 c/ml | 3 | 0 | 0 | 3 | 3 | 100 |
| MERS-coronavirus | 1.00E+04 TCID50/ml | 3 | 0 | 0 | 3 | 3 | 100 |
| Adenovirus | 1.00E+05 TCID50/ml | 3 | 0 | 0 | 3 | 3 | 100 |
| Human Metapneumovirus (hMPV) | 1.00E+06 TCID50/ml | 3 | 0 | 0 | 3 | 3 | 100 |
| Parainfluenza virus 1 | 1.00E+05 TCID50/ml | 3 | 0 | 0 | 3 | 3 | 100 |
| Parainfluenza virus 2 | 1.00E+05 TCID50/ml | 3 | 0 | 0 | 3 | 3 | 100 |
| Parainfluenza virus 3 | 1.00E+05 TCID50/ml | 3 | 0 | 0 | 3 | 3 | 100 |
| Parainfluenza virus 4 | 1.00E+03 TCID50/ml | 3 | 0 | 0 | 3 | 3 | 100 |
| Influenza A (H3N2) | 1.00E+05 TCID50/ml | 3 | 0 | 0 | 3 | 3 | 100 |

|  |  | SARS-CoV-2<br>Unspiked |  |  | SARS-CoV-2<br>Spiked <sup>a</sup> |  |  |
| --- | --- | --- | --- | --- | --- | --- | --- |
| Microorganism | Concentration | N Tested | N<br>Detected | %<br>Detected | N<br>Tested | N<br>Detected | %<br>Detected |
| Influenza B | 2.00E+03<br>TCID50/ml | 3 | 0 | 0 | 3 | 3 | 100 |
| Enterovirus (e.g. EV68) | 1.00E+05<br>TCID50/ml | 3 | 0 | 0 | 3 | 3 | 100 |
| Respiratory syncytial<br>virus | 1.00E+05<br>TCID50/ml | 3 | 0 | 0 | 3 | 3 | 100 |
| Rhinovirus | 1.00E+04<br>TCID50/ml | 3 | 0 | 0 | 3 | 3 | 100 |
| *Chlamydia pneumonia* | 1.00E+06<br>IFU/ml | 3 | 0 | 0 | 3 | 3 | 100 |
| *Haemophilus influenzae* | 1.00E+06<br>CFU/ml | 3 | 0 | 0 | 3 | 3 | 100 |
| *Legionella pneumophila* | 1.00E+06<br>CFU/ml | 3 | 0 | 0 | 3 | 3 | 100 |
| *Mycobacterium*<br>*tuberculosis* | 1.00E+06<br>TCID50/ml | 3 | 0 | 0 | 3 | 3 | 100 |
| *Streptococcus*<br>*pneumonia* | 1.00E+06<br>CFU/ml | 3 | 0 | 0 | 3 | 3 | 100 |
| *Streptococcus pyogenes* | 1.00E+06<br>CFU/ml | 3 | 0 | 0 | 3 | 3 | 100 |
| *Bordetella pertussis* | 1.00E+06<br>CFU/ml | 3 | 0 | 0 | 3 | 3 | 100 |
| *Mycoplasma*<br>*pneumoniae* | 1.00E+06<br>CFU/ml | 3 | 0 | 0 | 3 | 3 | 100 |
| *Pneumocystis jirovecii* | 1.00E+06<br>nuc/ml | 3 | 0 | 0 | 3 | 3 | 100 |
| *Candida albicans* | 1.00E+06<br>CFU/ml | 3 | 0 | 0 | 3 | 3 | 100 |
| *Pseudomonas*<br>*aeruginosa* | 1.00E+06<br>CFU/ml | 3 | 0 | 0 | 3 | 3 | 100 |
| Microorganism | Concentration | N Tested | N<br>Detected | %<br>Detected | N<br>Tested | N<br>Detected | %<br>Detected |
| *Staphylococcus epidermidis* | 1.00E+06<br>CFU/ml | 3 | 0 | 0 | 3 | 3 | 100 |
| *Streptococcus salivarius* | 1.00E+06<br>CFU/ml | 3 | 0 | 0 | 3 | 3 | 100 |
| Negative clinical NP<br>swab specimens (N=30<sup>b</sup>) | N/A | 90 | 0 | 0 | 90 | 90 | 100 |

<sup>a</sup>SARS-CoV-2 inactivated cultured virus spiked at 0.03 TCID<sub>50</sub>/ml (3x LoD)
<sup>b</sup>Each NP swab specimen was tested in triplicate for a total of 90 replicates in the absence and presence of spiked SARS-CoV-2 inactivated virus.

